# Leveraging Large Language Models to Map Triggers of Contamination-Related Obsessive-Compulsive Symptoms

**DOI:** 10.1101/2025.05.15.25327706

**Authors:** Dorothée Bentz, Dirk U. Wulff

## Abstract

Recent advancements in natural language processing (NLP) and large language models (LLMs) offer new avenues for exploring previously under-researched areas in mental health. Their capacity to automatically and meaningfully analyze large-scale text data makes them particularly valuable for studying highly individualized phenomena with clinical relevance, such as triggers of obsessive-compulsive symptoms (OCS), where pattern identification is often challenging. To address this gap, we surveyed 1,495 individuals from the general population about contamination-related obsessive–compulsive symptoms (C-OCS), as well as their triggers and corresponding intensity. Using LLM-based embeddings, we generated a map of key trigger categories for C-OCS, revealing their diversity across ecological domains and varying degrees of semantic similarity. Monte Carlo simulations further showed that individuals frequently reported semantically similar trigger pairs that differed in intensity. These findings provide a basis for further investigations into associative learning processes at the categorial and semantic levels. Such research in turn may enhance understanding of mechanisms involved in the development, maintenance, and treatment of obsessive-compulsive disorders, and may inform novel therapeutic approaches.

## Main

Rapid advances in automatic text data processing, particularly in natural language processing (NLP) and large language models (LLM), have the potential to bridge knowledge gaps in understanding mental disorders. NLP and LLM excel at analyzing large volumes of text data in a meaningful and Correspondence concerning this article should be addressed to Dorothée Bentz, Clinical Psychology and Translational Psychotherapy Research, Faculty of Psychology, University of Basel, Missionsstrasse 62A, 4055 Basel, Switzerland, and Dirk U. Wulff, Center for Adaptive Rationality, Max Planck Institute for Human Development, Lentzeallee 94, 14195 Berlin, automated manner (Chandran et al., 2019), and the growing availability of open-source LLMs further bolsters such research by enhancing transparency and reproducibility, which are crucial especially when investigating sensitive topics in mental health (Binz et al., 2025; Hussain et al., 2024; Wulff et al., 2024). This allows large populations to be surveyed on clinically relevant topics using open-text formats and to analyze their responses efficiently (Feuerriegel et al., 2025; Hussain et al., 2024). This approach is particularly useful for studying clinical phenomena that are both widespread and highly idiosyncratic, such as triggers of compulsive behaviors and obsessive thoughts related to contamination fears.

Contamination-related obsessive–compulsive symptoms (C-OCS) are common in the general population and are central to approximately half of the 2 to 3 percent of individuals with lifetime obsessive–compulsive disorder (OCD). In clinical presentations, contamination fears frequently lead to excessive and maladaptive washing or cleaning rituals, referred to as contamination-related OCD (C-OCD) (APA, 2013; Karno et al., 1988; Rasmussen & Eisen, 1992). Contemporary dimensional models of psychopathology conceptualize mental disorders like OCD as varying in severity along continuous symptom dimensions instead of representing discrete categories (Eaton et al., 2023). Consistent with this conceptualization, there is substantial evidence that C-OCS and C-OCD reflect quantitative differences along a shared continuum, rather than qualitatively distinct phenomena (Abramowitz et al., 2014; Adam et al., 2012; Angst et al., 2004; Haslam et al., 2005; Mataix-Cols et al., 2003; Mataix-Cols et al., 2005).

Individuals with C-OCS typically respond to a range of distinct stimuli (also called triggers), such as specific objects (e.g., toilets or garbage bins) or situations (e.g., shaking hands or using public transportation) (APA, 2013). However, comprehensive studies that systematically examine the nature and prevalence of these triggers are lacking. The existing knowledge comes largely from anecdotal reports from patients and OCD experts (Cullen et al., 2021; Mataix-Cols et al., 2009; Rachman, 2004; Simon et al., 2012; Sousa et al., 2024), as well as from symptom provocation studies in individuals with OCS, which have assessed C-OCS triggers based on dimensions derived theoretically, such as fear or disgust (Cullen et al., 2021; Mataix-Cols et al., 2009; Simonet al., 2012; Sousa et al., 2024). Consequently, general data on the frequency of specific triggers in both clinical and nonclinical populations remain scarce, as well as more detailed information on demographic patterns, such as whether certain triggers are more common in one gender or age group.

The lack of a comprehensive empirical understanding of triggers and their semantic relationships stands in stark contrast to their central role in exposure with response prevention (ERP), the first-line psychotherapeutic treatment for OCD, where individuals systematically expose themselves to these triggers (for Mental Health (UK). Leicester (UK): British Psychological Society (UK), 2006). In particular, clarifying the semantic relationships between triggers could help overcome a major limitation of all exposure-based therapies: success with one trigger does not always generalize to others (Jacoby & Abramowitz, 2016; Kodzaga et al., 2025). This limitation is particularly important because associative learning processes, which are considered critical for both OCD symptomatology and ERP efficacy, are highly dependent on trigger characteristics (Kodzaga et al., 2025; Pittig et al., 2016). In fear learning, associations between one stimulus and another are transferred based on their perceived similarity (Hall, 1996; Shepard, 1987, 2004), whereas generalization occurs only to a limited extent or not at all in extinction (Kodzaga et al., 2025; Vervliet et al., 2010; Vervliet et al., 2005).

In the context of OCS, only a few studies have investigated the generalization of triggers (Cooper & Dunsmoor, 2021). In particular, there is a lack of studies examining generalization at the conceptual level (Dunsmoor et al., 2011; Vervoort et al., 2014) and at the semantic-relational level (Boyle et al., 2016). Yet, it might be, in particular, these conditioning paradigms that assess generalization along conceptual or semantic dimensions rather than only along the perceptual dimension that may best capture the often complex and abstract nature of triggers for OCS (Cooper & Dunsmoor, 2021). A key reason for the scarcity of research in this area might be the limited understanding of the semantic relationships of the OCS triggers. A basis for this research is an expansion of the currently limited understanding of the semantic relationships of OCS triggers. Expanding this line of research could also enable targeted associative learning experiments examining dysfunctional cognition, such as threat overestimation, which is considered relevant to OCD symptomatology (Hezel & McNally, 2016).

Taken together, advancing research on extinction generalization - and potentially improving the treatment of OCS - requires a comprehensive understanding of the nature of OCS triggers and their semantic relationships. This study aims to address this gap by leveraging LLMs to map C-OCS triggers using a large dataset of free text responses from individuals in the general population who self-report to experience C-OCS. Specifically, we examine the frequency and content of various triggers, categorize them into distinct categories of triggers of C-OCS, and analyze their distribution between individuals with varying severity of C-OCS, as well as across different age and gender groups. In addition, we investigate the relationship between trigger intensity and semantic similarity to identify semantically similar stimuli with divergent trigger intensity. This approach lays the foundation for future research on associative learning in OCS and ERP, with the potential to inform novel intervention strategies.

## Methods

### Design, Setting, and Participants

The study utilized an anonymous online survey to investigate OCS triggers related to washing and contamination symptoms. Participation was offered as a follow-up to the Swiss Corona Stress Study (Survey 4) (Freytag et al., 2022), which was conducted between November 16 and 28, 2021, to assess the COVID-19 pandemic’s impact on mental wellbeing in Switzerland. Participants for the Swiss Corona Stress Study were recruited from all Swiss regions via media releases from the University of Basel, local newspapers, radio interviews, and social media. Inclusion criteria for the Swiss Corona Stress Study were Swiss residency, 14 years or older, and no participation in previous iterations of the study (Surveys 1-3) (de Quervain et al., 2020a, 2020b). All participants provided written informed consent prior to participation and received no monetary compensation. From the 11,167 individuals in the Swiss Corona Stress Study (Survey 4) (Freytag et al., 2022), 3,615 voluntarily proceeded to a survey on compulsions and obsessions. Of these, 1,213 participants (80.6% female, mean age 29.8 years) reported personal triggers for washing/contamination symptoms and rated the intensity of these C-OCS triggers.

The data analyzed in this study were collected as part of the Swiss Corona Stress Study, which consisted of an anonymous online survey. In accordance with Swiss legislation—specifically the Swiss Human Research Act (HRA, Art. 2, para. 2, letter c)—research conducted with anonymized health-related data is explicitly exempt from the scope of this law and does not require approval by an ethics committee; therefore, no formal ethics approval or exemption was required or obtained. Our study complied fully with applicable Swiss data protection regulations and ethical standards for research involving anonymized data.

### Procedure

The anonymous online survey was structured in two parts. The first part comprised the Swiss Corona Stress Study (Survey 4) (Freytag et al., 2022), and the second was an optional, independent survey focusing on compulsions, obsessions, and triggers within the washing/contamination dimension of OCS. The first part took approximately 15 min-utes to complete, and the second part took about 5 minutes. The participants accessed the survey via the website www.coronastress.ch, available in German, French, and Italian. The survey was implemented using SoSci Survey software (Leiner, 2019), which recorded the participation date but not IP addresses or timestamps. Completion required responses to all items.

Following general study information and informed consent, the Swiss Corona Stress Study collected self-report sociodemographic data (gender, age, nationality, education, profession), self-declared psychiatric conditions, and information on stress and behavioral changes during the pandemic. Upon completing the first part, participants were invited to continue with the survey on compulsions and obsessions. These were defined as: “compulsions and obsessions can be activities or thoughts that you repeat or think about over and over again, e.g., washing very often, checking things, or counting, even if you do not want to or if they seem excessive or irrational to you.”

First, current obsessive-compulsive symptoms in the washing/contamination dimension were assessed using the washing subscale of the Obsessive-Compulsive Inventory-Revised (OCI-R). Second, participants were asked to list their “most common triggers for intrusive and distressing thoughts about germs and contamination and/or recurring behaviors or rituals to prevent contamination (e.g., washing, cleaning, showering)” in a free-text format and to indicate the intensity of distress caused by each trigger. At the study’s conclusion, participants received automated stress management recommendations based on their responses in the initial survey part (as part of the Swiss Corona Stress Study, Survey 4). Additionally, general information on OCS, treatment options, and contact details for professional support were provided.

### Measures

Contamination-related obsessive-compulsive symptoms (C-OCS) were assessed using the washing subscale of the Obsessive-Compulsive Inventory–Revised (OCI-R; items 5, 11, and 17), taken from the validated German (Gönner et al., 2007), French (Zermatten et al., 2006), and Italian (Marchetti et al., 2010) versions. The OCI-R is an 18-item self-report questionnaire assessing six dimensions of obsessive–compulsive symptoms (checking, washing, ordering, obsessing, neutralizing, and hoarding) using a 5-point response scale from 0 (not at all) to 4 (extremely). This yields a total score ranging from 0 to 72, with subscale scores ranging from 0 to 12, where higher scores indicate greater symptom severity. For the present study, only the washing subscale was administered to provide a brief and symptom-specific assessment of C-OCS. Prior research indicates that the washing subscale demonstrates high diagnostic accuracy, with a cut-off score >3 effectively distinguishing individuals with clinically significant washing compulsions from healthy controls (98 percent vs. 91 percent; Youden Index = 0.89) (REF: Gönner). Using the OCI-R subscale therefore allowed for efficient yet valid screening of C-OCS in our sample. Individual free-text responses regarding the “most common triggers” were used to identify C-OCS triggers. To prepare these responses for analysis, the first author manually translated the German, French, and Italian entries into English, with the second author validating these translations. The intensity of each trigger was determined from participants’ ratings of distress caused by their individual triggers, on a scale from 0 (none) to 10 (maximum).

### Creating the C-OCS trigger map

To create a map of trigger categories, we followed a multi-step process. First, we embedded the reported C-OCS triggers using a fine-tuned embedding model (specifically, dwulff/mpnet-cocs on Hugging Face) to capture their semantic organization, leveraging open-source platforms that promote accessibility and reproducibility in model sharing (Hussain et al., 2024). This model was fine-tuned using 20 thousand ratings generated by Llama-3.3-70b-Instruct Grattafiori et al., 2024. Llama was instructed as follows: “We have asked laypeople to name their C-OCS triggers. Your task is to evaluate on a scale from 0 to 100 whether two respondents named exactly the same trigger. Evaluate this in the context of contamination OCD. Briefly reason through your answer. Then return the answer as a number between 0 (fully different) and 100 (fully same). Strictly use the format Answer=[evaluation].” Finetuning was used to adapt the embedding model to the domain of C-OCS triggers, which is important given that many trigger responses include many everyday objects and activities whose dominant meaning is unrelated to C-OCS or OCD in general.

Based on this model, C-OCS triggers were then grouped into semantically cohesive groups. The average embedding of each group was subsequently projected into a two-dimensional map using the PaCMAP dimensionality reduction algorithm. Each group was labeled using Llama-3.3-70b-Instruct, using the following prompt: “Your task is to provide a short label (1 to 3 words) for the following list of C-OCD trigger responses: List: {trigger_text}. The label should be highly specific, closely capture the elements in the list, and should make sense as a trigger without adding meaning. The best label is often the most frequent element, potentially shortened. Only return the label!” The average similarity between labels and associations is.87, vastly exceeding the average similarity across all trigger responses (.26), supporting the suitability of LLM-generated cluster labels.

Finally, this map was partitioned into twelve clusters using hierarchical clustering. The number of clusters was selected manually based on the interpretability of the resulting cluster solution (Haslbeck & Wulff, 2020; Hennig, 2015, cf.) aiming to identify meaningful trigger categories.

### Measuring C-OCS trigger co-occurrence propensity

To identify trigger co-occurrences that exceed chance levels, we employed a simulation-based approach. Specifically, we simulated ten million synthetic respondents. These simulations used the inter-participant distributions of response numbers and overall trigger frequencies, assuming that these two distributions were independent. Based on this simulated data, we determined the likelihoods of trigger co-occurrences within the responses of the synthetic respondents. This allowed us to establish random expectations (p) for each trigger pair. We then used these random expectations to calculate a z-scaled co-occurrence score, applying a continuity correction for the binomial distribution. The formula used was 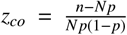, where *n* is the number of observed co-occurrences and *N* is the total number of possible pairs given the empirical distribution of response numbers.

We compared the co-occurrence scores for each pair with the intensity delta, computed as the absolute difference between the average intensities *I*_*i*_ and *I* _*j*_ for the two triggers *i* and *j* in the pair, i.e., Intensity delta = |*I*_*i*_ − *I* _*j*_|.

## Results

Our results are based on self-reported C-OCS symptom severity measured with the OCI-R washing subscale, as well as individual self-reported C-OCS triggers and their individually assessed trigger intensity. Our analysis includes 1,213 individuals with a score greater than zero on the OCI-R washing subscale, out of 1,399 participants who reported C-OCS triggers. In total, they identified 3,508 triggers. The results are structured as follows. First, we present a map of C-OCS trigger categories derived from LLM embeddings and subsequent cluster analysis, addressing our aim to identify and categorize the individually self-reported triggers. Second, we examine these categories in terms of retrieval order and how their prominence relates to C-OCS symptom severity, age, and gender to document key individual differences as outlined in our study objectives. Third, we analyze how semantic similarity influences trigger co-occurrences within individuals and variations in trigger intensity, exploring a potential pathway to enhance understanding of mechanisms relevant to treatment.

### Mapping C-OCS triggers

To create a map of trigger categories, we first embedded the triggers using a fine-tuned embedding model to understand the semantic organization of C-OCS triggers (see huggingface.co/dwulff/mpnet-cocs). We recruited a finetuned model to emphasize the OCD-related meaning of triggers, which are mostly everyday objects and activities. Without domain adaptation, the embedding model would not give appropriate weight to the unique context of this investigation. Based on this model, we then grouped C-OCS triggers into semantically cohesive groups and projected the average embedding of these groups into a two-dimensional map using PaCMAP. Finally, we partitioned the map into twelve clusters using hierarchical clustering, creating highly interpretable trigger categories. We manually selected the number of clusters based on the interpretability of the cluster solution.

In 1A, the C-OCS trigger categories are represented by different colored points and shaded areas on the map. Analyzing the twelve clusters, we identified seven as containing cohesive groups of C-OCS triggers. The remaining five clusters contained groups of “Other” responses, consisting of idiosyncratic C-OCS triggers, responses reflecting current social topics at the time of data collection (e.g., “Vaccination” or “Antivax lies”), or non-informative responses (e.g., “None” or “Nothing”). We focus our analysis on the seven cohesive groups of C-OCS triggers to avoid placing too much weight on idiosyncratic or potentially invalid responses, which are common in free-text responses and would otherwise distort our analysis. The excluded responses accounted for a small minority of responses (10.9%; see 1B) and were retrieved considerably later than the included responses (see 1C), which may suggest that they are produced based on associative processes rather than representing key C-OCS triggers.

Considering valid responses, the largest C-OCS trigger cluster, labeled *Surfaces*, accounts for 23.5% of responses and contains triggers such as “Door handle,” “Elevator buttons,” and “Shopping cart.” The second-largest cluster, labeled *Sickness* (18.5% of responses), contains triggers such as “Coughing/Sneezing,” “People without masks,” and “Corona.” The third-largest cluster, labeled *Proximity* (16.2%), contains triggers such as “Public transport,” “Shaking hands,” and “Crowd.” The fourth-largest cluster, labeled *Grime* (10.8%), contains triggers such as “Public toilets,” “Dirt,” and “Body odor.” The fifth-largest cluster, labeled *Cleaning* (10.4%), contains triggers such as “Washing hands,” “Showering,” and “Cleaning.” The sixth-largest cluster, labeled *Places* (7.9%), contains triggers such as “Shopping,” “Stores,” and “Restaurant.” Finally, the seventh-largest cluster, labeled *Food* (7.9%), contains triggers such as “Fruit and vegetables,” “Food packaging,” and “Eating.”

Taken together, our LLM-driven approach successfully mapped the diverse landscape of C-OCS triggers, revealing their semantic organization and providing a foundational, data-driven understanding of the key categories that provoke C-OCS, which has previously been reliant on anecdotal reports.

### Individual differences in C-OCS trigger importance

Our second goal was to understand individual differences in C-OCS trigger importance. We analyzed the relative response frequencies by the OCI-R washing score, age, and gender. Figure 2 shows the result for the seven valid categories.

**Figure 1.**
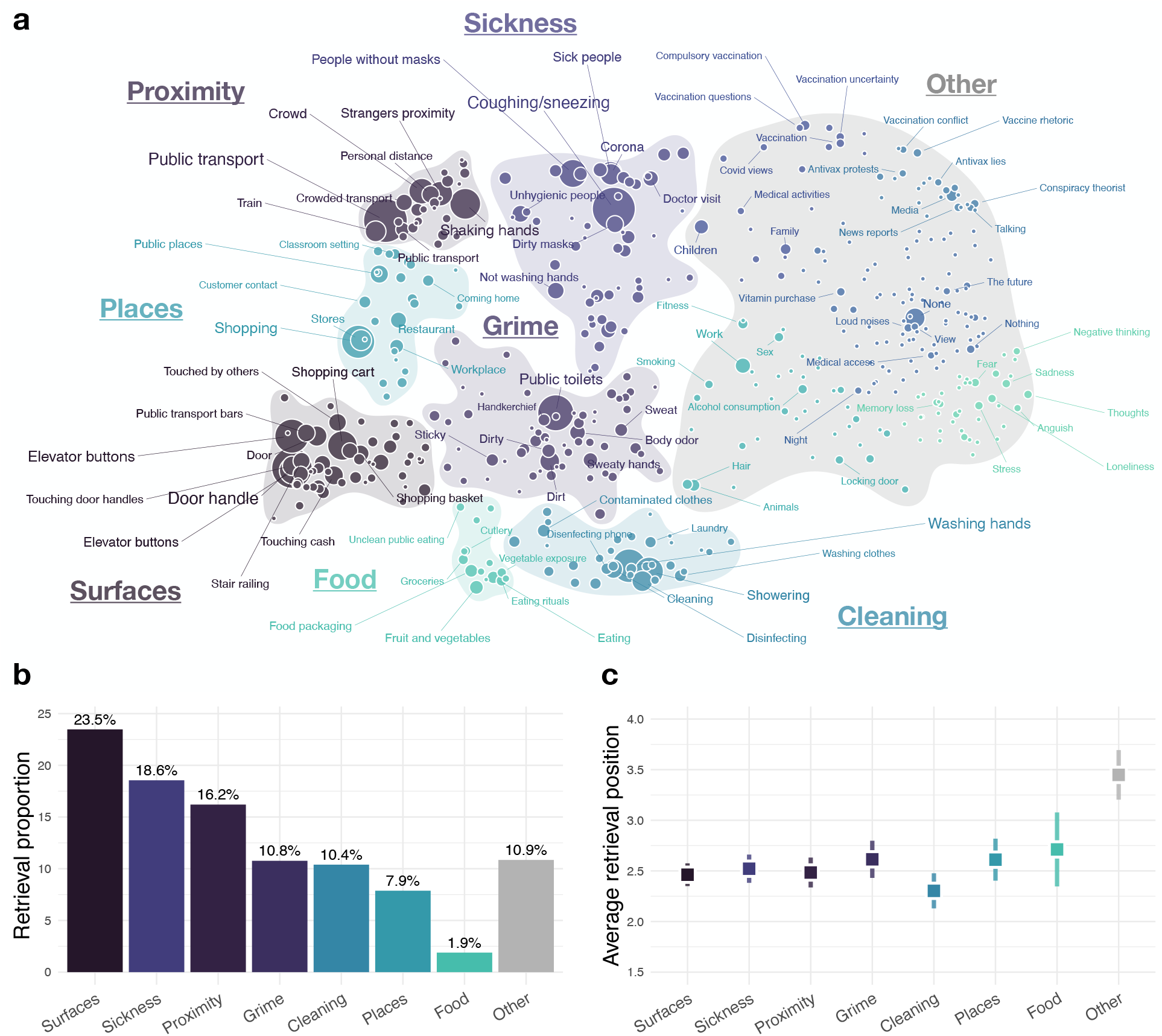
Map of C-OCS triggers. Panel A shows the C-OCS trigger map, generated using the PaCMAP dimensionality reduction algorithm. We identified twelve C-OCS trigger categories, each represented by a different color, including seven prominent categories identified as valid C-OCS triggers. Labels were assigned to best represent the semantic content of each trigger cluster. Panels B and C show, respectively, the proportion and average position of participant responses belonging to each cluster.

**Figure 2.**
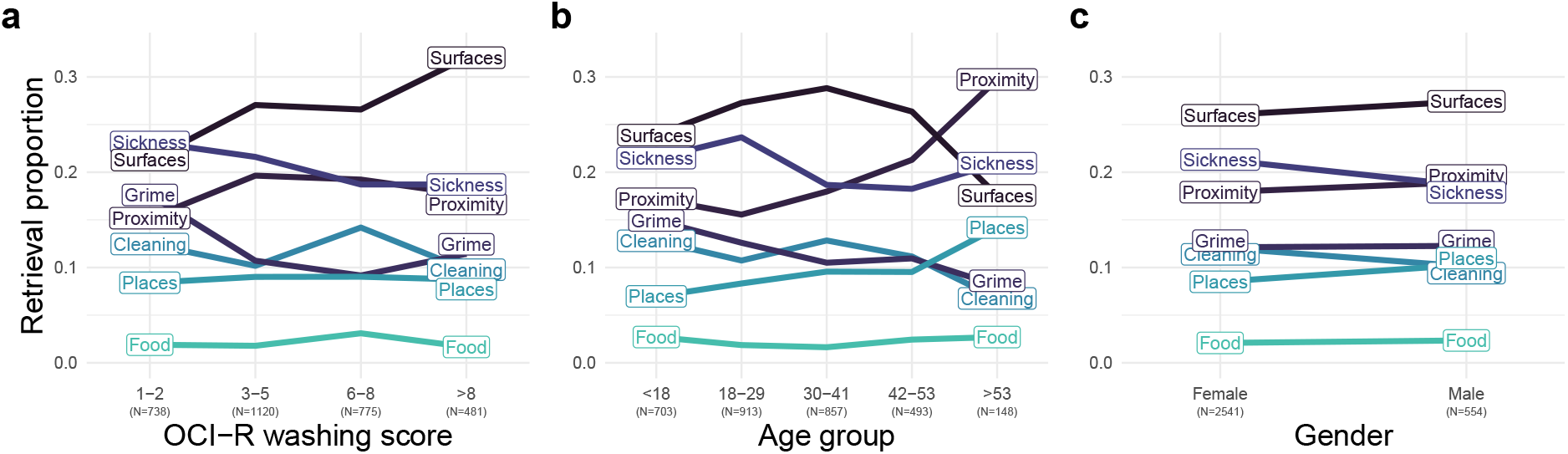
OCI−R washing score Age group Gender C-OCS trigger moderators. The figure shows the relative frequency of triggers from the 7 trigger categories as a function of C-OCS symptom severity (a), age (b), and gender (c).

Overall, the importance of categories was relatively stable across the three individual differences variables, as indicated by rank-correlations across levels of *r* =.91 (OCI-R washing score), *r* =.87 (age group), and *r* =.93 (gender). However, some notable deviations emerged. Analyzing the responses from each cluster as a function of all three individual variables simultaneously, we found that *Surfaces* was more prominent among high compared to low-symptom individuals, while the opposite was true for *Sickness* and *Grime*. Furthermore, *Proximity* and *Places* were more prominent among older as compared to younger adults, whereas the opposite was true for *Grime*. No significant differences were observed for gender.

These findings suggest that while the key C-OCS trigger categories identified are broadly relevant, their prominence can exhibit subtle yet potentially meaningful variations related to symptom severity and age. This nuanced understanding contributes to a more detailed picture of how C-OCS triggers manifest across different segments of the population experiencing these symptoms.

### Evaluating the potential of semantic trigger replacement

Our third goal was to evaluate whether semantic similarity between C-OCS triggers could be leveraged to improve ERP and potentially increase patients’ acceptance of the method. Specifically, we aimed to identify triggers that are semantically similar but frequently co-occur within the same individual, yet are rated as having different intensity levels. Such constellations may represent attractive targets for what we refer here to as semantic trigger replacement, namely selecting or sequencing trigger presentations based on their semantic similarity to other C-OCS triggers. This approach may allow clinicians to deliberately select semantically similar but less intense triggers to facilitate generalization to more intense triggers, thereby creating new therapeutic opportunities within the ERP framework.

To analyze the potential for semantic trigger replacement, we first developed a simulation-based approach to quantify trigger co-occurrence. We simulated ten million synthetic respondents under the assumption of trigger independence and then calculated the propensity by z-standardizing the empirical co-occurrence frequencies using the synthetic ones. This approach corrects for the effect of trigger frequency on co-occurrences, producing a cleaner measure of trigger cooccurrence (Goñi et al., 2011; Wulff et al., 2022).

Figure 3A shows the relationship between the trigger cooccurrence propensity for each trigger pair with a minimum co-occurrence frequency of five and the corresponding intensity delta, calculated as the absolute differences between the trigger intensity scores of the trigger pair. The labels show pairs that are high in either frequency, co-occurrence propensity, or intensity delta. The two variables show a mild positive relationship (*r* =.20), with higher co-occurrences going hand-in-hand with higher intensity deltas. This pattern implies that there exist trigger pairs that frequently co-occur but do not elicit C-OCS with the same intensity. For instance, “Showering” co-occurred frequently with both “Cleaning” and “Washing hands” but elicited substantially smaller intensity ratings. Such trigger pairs could represent effective candidates for semantic trigger replacement in ERP and open the possibility of selecting a less intense trigger as a starting point for ERP.

**Figure 3.**
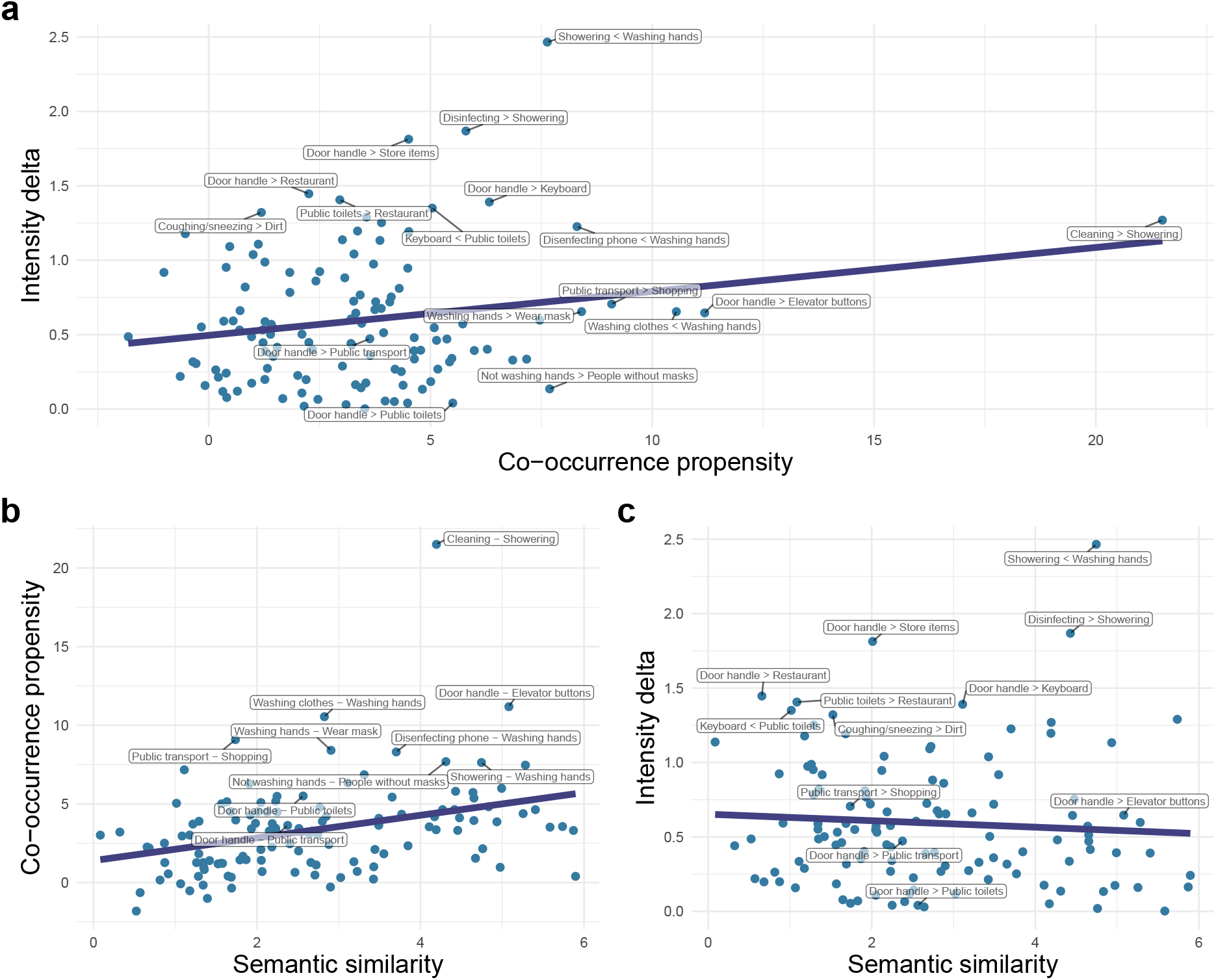
Potential for C-OCS trigger replacement. The figure shows the relationships between semantic trigger similarity, cooccurrence propensity, and intensity delta. Panel a shows the relationship between co-occurrence propensity and trigger intensity delta (difference in trigger strength among pairs). Panels b and c show the relationship of semantic similarity to co-occurrence propensity and intensity delta, respectively.

For semantic trigger replacement to be effective, semantic similarity must serve as a close proxy for trigger cooccurrences, but not for intensity. To evaluate this, we analyzed the corresponding correlations. As can be seen in Figure 3B-C, semantic similarity was positively related to co-occurrence propensity (*r* =.35, *CI*_95_ = [.18,.50], *t*_120_ = −4.1, *p* <.001) and not related to intensity delta (*r* =−.07, *CI*_95_ = [−.25,.11], *t*_120_ = −.77, *p* =.445), implying that high-similarity triggers are more frequently named by the same person than low-similarity triggers, while not significantly differing in intensity. This suggests that semantic similarity is a useful proxy for identifying conceptually related triggers that are likely to co-occur, even if they do not always elicit the same level of C-OCS.

All in all, these analyses demonstrate that semantically similar C-OCS triggers often co-occur within an individual’s reported triggers, yet can significantly differ in their reported intensity. This finding highlights the potential of using semantic similarity as an empirical guide to select and sequence stimuli in exposure therapy, thereby paving the way for more nuanced and potentially more effective treatment strategies aimed at improving extinction generalization for OCS.

## Discussion

Leveraging a large language model (LLM) to analyze free-text responses about triggers of C-OCS in the general population, our study demonstrates that these triggers—often considered highly idiosyncratic—can be meaningfully grouped into distinct categories. Specifically, our LLM-driven mapping and clustering identified seven primary C-OCS trigger categories: *Surfaces, Sickness, Proximity, Grime, Cleaning, Places*, and *Food*. These categories span various semantic domains and are linked to different ecolog-ical contexts of individuals. Unlike previous research that often relied on predefined stimuli rated for intensity (Mataix-Cols et al., 2009), our approach gathered free-text responses from individuals who self-report C-OCS about their personal triggers, which were then rated for intensity. Our findings extend descriptive work such as that of Rachman (2004), who identified heterogeneous contamination triggers and grouped them broadly. While some of our empirically identified categories show conceptual overlap with theirs (e.g., *Sickness* with diseases/germs, *Grime* with dirt/pollution), our study uniquely quantifies their prevalence and semantic organization based on a large dataset. The most frequently reported trigger categories in our sample were *Surfaces* (23.5%), followed by *Sickness* (18.5%) and *Proximity* (16.2%), highlighting the key areas of concern for individuals with C-OCS.

Examining individual differences revealed a nuanced picture regarding the prominence of these trigger categories. The influence of the severity of the C-OCS, for example, showed that while the overall importance of the cluster rank was relatively stable, supporting a dimensional approach to OCS (Abramowitz et al., 2014; García-Soriano et al., 2011), some significant deviations emerged. *Surfaces* were more prominent for individuals with higher symptom severity, whereas *Sickness* and *Grime* were more prominent for those with lower severity, suggesting that the salience of certain trigger categories may shift with symptom severity. Agerelated variations were also notable: triggers related to *Proximity* and *Places* were more prominent among older adults, while *Grime* was more so among younger adults. These age-based differences might reflect genuine developmental shifts in C-OCS trigger prominence or could be partially influenced by the data collection period during the COVID-19 pandemic, which heightened concerns about public spaces and proximity, particularly for older individuals (Andrighetto et al., 2024). In contrast to some existing literature suggesting gender differences in C-OCS categories (Bogetto et al., 1999; Labad et al., 2008), our analysis did not reveal significant differences in the prominence of the identified trigger categories between genders. Together, these findings underscore that while key trigger categories are broadly relevant, their specific importance can vary across different segments of the population experiencing C-OCS, offering potential avenues for tailoring therapeutic interventions.

A potentially clinically significant finding is that semantically similar triggers, which frequently co-occur within an individual’s reported triggers, can differ notably in their perceived intensity. Our analysis showed a positive relationship between semantic similarity and co-occurrence propensity, but no substantial relationship between semantic similarity and the difference in trigger intensity (intensity delta). This suggests that individuals often report multiple semantically related triggers, but these triggers do not necessarily elicit the same level of distress. This insight would have direct implications for exposure with response prevention (ERP): if future research confirms a robust generalization of extinction learning between semantically similar stimuli within the same or even to adjacent dimensions, as suggested by studies on associative learning in OCD (Cooper & Dunsmoor, 2021). Then lower intensity triggers could potentially be used during ERP to facilitate generalization to related, higher intensity triggers. Such an approach would potentially enhance treatment tolerability and reduce dropout rates, as exposure to highly intense triggers is a known barrier to ERP engagement and completion (Ong et al., 2016; Öst et al., 2015; Rosa-Alcázar et al., 2008).

The methodological approach employed in this study, particularly its reliance on open LLM technologies, holds considerable promise for broader applications. The use of finetuned models available through open-source platforms reduces barriers to entry and encourages the adaptation of these powerful tools for various clinical research questions (Hussain et al., 2024; Wulff et al., 2024). The use of LLMs to systematically analyze and map idiosyncratic, free-text descriptions of symptom triggers is not limited to C-OCS (Aeschbach et al., 2025). This data-driven technique could be readily adapted to explore the trigger landscapes of other OCS dimensions, such as checking compulsions (e.g., identifying common fears or behavior excesses), or symmetry and ordering concerns. Beyond OCS, this methodology could be invaluable in understanding triggers and expressions of symptoms in other mental health conditions such as PTSD (characterizing trauma-related signals), anxiety disorders (mapping phobic stimuli or content of worries), or even depressive disorders (identifying patterns in automatic negative thoughts). Furthermore, in general medicine, analyzing patient-reported outcomes or qualitative descriptions of symptoms through LLMs could help in understanding complex, multifaceted conditions where individual experiences vary widely, such as chronic pain or autoimmune diseases, thereby fostering a more nuanced, patient-centered approach to clinical research and practice.

Our LLM-based approach provides a more fine-grained, data-driven understanding of the semantic landscape of C-OCS triggers. This provides crucial insights that can inform the refinement of treatment strategies, particularly ERP. By moving beyond predefined or purely anecdotal categorizations to a comprehensive semantic mapping, we extend current knowledge and enable further research into the generalization of extinction. This is in line with a recent systematic review on extinction generalization following exposure in anxiety disorders, which emphasizes that clinical progress depends on a more nuanced understanding of stimulus complexity and the factors driving generalization (Kodzaga et al., 2025). Understanding these semantic relationships is vital for advancing research into the associative learning mechanisms that underpin successful OCS treatment and for guiding the development of more personalized and potentially more effective therapeutic interventions. At the same time, we emphasize that the clinical implications of our findings remain preliminary. An essential next step will be to validate this semantic structure in clinical populations and to test its relevance in controlled conditioning paradigms.

We have demonstrated that recent advances in natural language processing, specifically large language models, can make a significant contribution to psychopathology research, particularly in understanding obsessive-compulsive symptoms. Our analysis revealed that C-OCS triggers, often perceived as highly idiosyncratic, can be systematically mapped and grouped into meaningful semantic categories based on individuals’ free-text descriptions. These findings not only provide a richer, empirically grounded taxonomy of C-OCS triggers but also enable further investigation into their role in associative learning experiments, ripe for validation in clinical populations. Ultimately, this data-driven approach to understanding symptom triggers contributes to enhancing the efficacy and personalization of OCD treatment and offers a versatile methodology for exploring similar phenomena across a range of psychological and medical conditions.

### Limitations

We would like to point out three important limitations. First, our sample may not be fully representative. Recruitment was conducted within a study on stress and behavioral changes during the COVID-19 pandemic, which might have introduced a selection bias towards individuals more attuned to mental health issues or experiencing higher stress. Data collection during the pandemic may have also influenced the prominence of certain contamination triggers, particularly those related to infectious diseases (e.g., the *Sickness* cluster). Although participants were asked to report triggers beyond official hygiene measures, the prevailing context may have heightened awareness of them. This aligns with observed increases in contamination-related compulsive symptoms during that period (Otte et al., 2025). Therefore, while these findings are highly relevant, replication in diverse representative samples and in a post-pandemic context would strengthen their generalizability. Second, our sample is drawn from a mostly non-clinical population, which may differ from a clinical population in the distribution of C-OCS triggers. Furthermore, we base our inclusion criterion on the OCI-R washing subscale, an efficient screening instrument that, however, lacks normative data for use in isolation. Consequently, it is currently unclear how well our results generalize to a clinical population. Future studies should evaluate the distribution of triggers in clinical samples, ideally using additional diagnostic measures, such as clinical interviews, to further substantiate symptom severity. Third, responses likely have been affected by associative retrieval processes (Aeschbach et al., 2025; Wulff & Mata, 2022). Listing triggers of obsessive compulsive symptoms necessarily relies on memory retrieval, which is known to be affected by various cognitive factors (Wulff et al., 2019). Based on this reason, we have excluded several minor clusters from the analysis. While this decision was also informed by retrieval position and null responses (e.g., “none”), we cannot rule out that many responses grouped under *Other* could be genuine OCS triggers. Conversely, we also cannot fully rule out that the other categories are unaffected by associative processes. This could, for instance, explain the importance of the *Cleaning* category, which may have resulted from conflating triggers with associated consequences. That said, past work sug-gests that cleaning itself may trigger symptoms. For instance, (Zisler et al., 2024) found that 70% of individuals with OCD experienced obsessions and compulsions when cleaning the apartment, suggesting that cleaning is a valid trigger category.

## Declarations

### Data availability statement

Data and materials are available at https://osf.io/edyg5/.

### Code availability statement

Data and materials are available at https://osf.io/edyg5/.

## Acknowledgments

DB acknowledges that data collection was carried out within the framework of the Swiss Corona Study, supported by funding from the Transfaculty Research Platform for Molecular and Cognitive Neurosciences, University of Basel, and funds from the research fund of the University of Basel granted to DB. The authors thank Nathalie Schicktanz for her contributions to survey implementation and Thomas Schlitt for data management. DUW acknowledges funding from the German Research Foundation (https://gepris.dfg.de/gepris/projekt/546419617?language=en).

## Author information

Dorothée Bentz, Department of Biomedicine, Division of Cognitive Neuroscience and Faculty of Psychology, Clinical Psychology, and Translational Psychotherapy Research, University of Basel, Missionsstrasse 62A, 4055 Basel, Switzerland Dirk U. Wulff, Center for Adaptive Rationality, Max Planck Institute for Human Development, 14195 Berlin, and Cognitive and Decision Sciences, Faculty of Psychology, University of Basel, Missionsstrasse 64A, 4055 Basel, Switzerland.

## Contributions

DB and DUW designed the study. DB supervised survey implementation and data collection. DUW analyzed and visualized the data. DB and DUW wrote the manuscript.

